# Association of Systemic Immune-Inflammation Index with Cognitive Impairment and Magnetic Resonance Imaging Markers in Patients with CADASIL

**DOI:** 10.64898/2026.01.13.26344080

**Authors:** Jinghua Peng, Hongri Zhang, Fangwei Hu, Guangpeng Leng, Zhe Peng, Shuyan Xu, Tao Yang, Wenxin Qiu, Chaohan Chen, Bin Cai

## Abstract

**Background:** Cerebral autosomal dominant arteriopathy with subcortical infarcts and leukoencephalopathy (CADASIL) is a monogenic hereditary cerebral small vessel disease (CSVD). Existing studies have confirmed that it is caused by mutations in the *NOTCH3* gene, but the specific mechanisms underlying its pathogenesis and progression remain elusive. Existing research indicates that inflammation plays a critical role in the development and progression of CSVD. The Systemic Immune-Inflammation Index (SII) has revealed to be a reliable new marker to assess immune status and inflammatory response intensity. This study reveals the relationship of SII with cognitive impairment and magnetic resonance imaging (MRI) markers of CSVD in patients with CADASIL.

**Methods:** This cross-sectional investigate included patients diagnosed with CADASIL who had confirmed *NOTCH3* gene mutations and complete clinical data. Cognitive function in patients with CADASIL was appraised by the Mini-Mental State Examination (MMSE). SII is obtained by calculating the number of platelets, neutrophils and lymphocytes in blood routine examination. Summary SVD score and imaging markers of CSVD, including cerebral microbleeds, lacunae, white enlarged perivascular space and matter hyperintensity were evaluated based on magnetic resonance imaging. The association between cognitive impairment and SII and MRI markers in CADASIL were evaluated using logistic regression models and Spearman correlation.

**Results:** At baseline, A total of 96 Patients with CADASIL were enrolled in this cross-sectional study. the median age of patients with CADASIL was 59.00 (interquartile range 52.25-66.75) years, and 58.3% of patients were male. The correlation analysis results indicate that the SII level was negatively correlated with MMSE scores in patients with CADASIL (rs=-0.336, *P* <0.001). An elevated SII was statistically significantly linked with the risk of cognitive impairment (Q4 vs. Q1: OR 5.230, 95% CI 1.040-26.297; *P*=0.045) after adjusting for age, sex and education. In contrast, there was no considerable difference between SII and summary SVD score or MRI imaging markers.

**Conclusions:** Elevated SII was linked with cognitive impairment in CADASIL patients. Nevertheless, there were no significant differences between SII and summary SVD score or MRI imaging markers.

## 1 Introduction

CSVD contributes to 25% of stroke and 45% of all cognitive impairment or dementia^(1–5)^. CADASIL is the most common hereditary CSVD. The causative gene for CADASIL is the notch receptor 3 gene (NO*TCH3*, OMIM 600276) which locate on chromosome 19p13.12. Granular osmiophilic material (GOM) has been used as a diagnostic markers which was observed in vascular smooth muscle cells (VSMCs) both in the brain and skin^(6, 7)^. Typical CADASIL symptoms include recurrent ischemic strokes or transient ischemic attacks (TIAs), migraine, cognitive impairment and psychiatric disturbances. The main characteristic MRI presentations in CADASIL patients include lacunes, white matter hyperintensity (WMH), enlarged perivascular space (EPVS) and cerebral microbleeds (CMBs)^(6–11)^.

Although many studies have been conducted on the pathogenesis of CADASIL, and the specific mechanism is not completely clear. Recent studies have shown that inflammation contributed significantly to the pathophysiological mechanisms in the CSVD^(5,7)^. Existing research has confirmed that systemic inflammation influenced the occurrence and progression of CSVD, SII (platelet count × neutrophil count/lymphocyte count) has been recommended as a novel biomarker of systemic inflammatory response which is easily accessible and can assess the immune and inflammatory status. SII was associated with cerebral blood flow autoregulation, endothelial dysfunction, and blood–brain barrier permeability^(12)^. Previous studies have revealed that SII was associated with patient outcomes with malignant tumors^(13)^, It may provide a new biomarker for prognosis in autoimmune diseases, cardiovascular and cerebrovascular^(14–18)^. In our study, was explored the association of SII with cognitive impairment as well as imaging markers and summary SVD score in a CADASIL cohort to identify new potential markers for predicting onset and progression in patients with CADASIL,which has not been explored.

## 2 Methods

### 2.1 Participants and Study design

Data for this cross-sectional study were obtained from a prospective CADASIL registry cohort, which has been detailed in our previous research^(19)^. All the participants included in the study were confirmed cysteine-altering variants in *NOTCH3* by Sanger sequencing or second-generation sequencing, and the clinical and neuroimaging of the participants were analysised. *NOTCH3* gene mutation was detected in 699 CSVD patients suspected of CADASIL, A total of 258 *NOTCH3* gene mutation patients were detected. Among them, 96 subjects with complete clinical data, laboratory tests data and MRI information were enrolled in this study. All patients were older than 18 years old. Patients who had contraindications to MRI, unable to complete clinical and imaging evaluation, complicated with cancer, active infection, immune system and blood system diseases, mental diseases, severe heart, liver and kidney diseases, cognitive impairment caused by other reasons, using immunosuppressants, recent history of trauma or surgery were excluded(Figure 1). Our study was approved by the Ethics Committee, and registered at ClinicalTrials. Gov, identifier: (NCT04318119, Registration Date03/21/2020).Informed consent was obtained from all participants.

**Figure 1.**
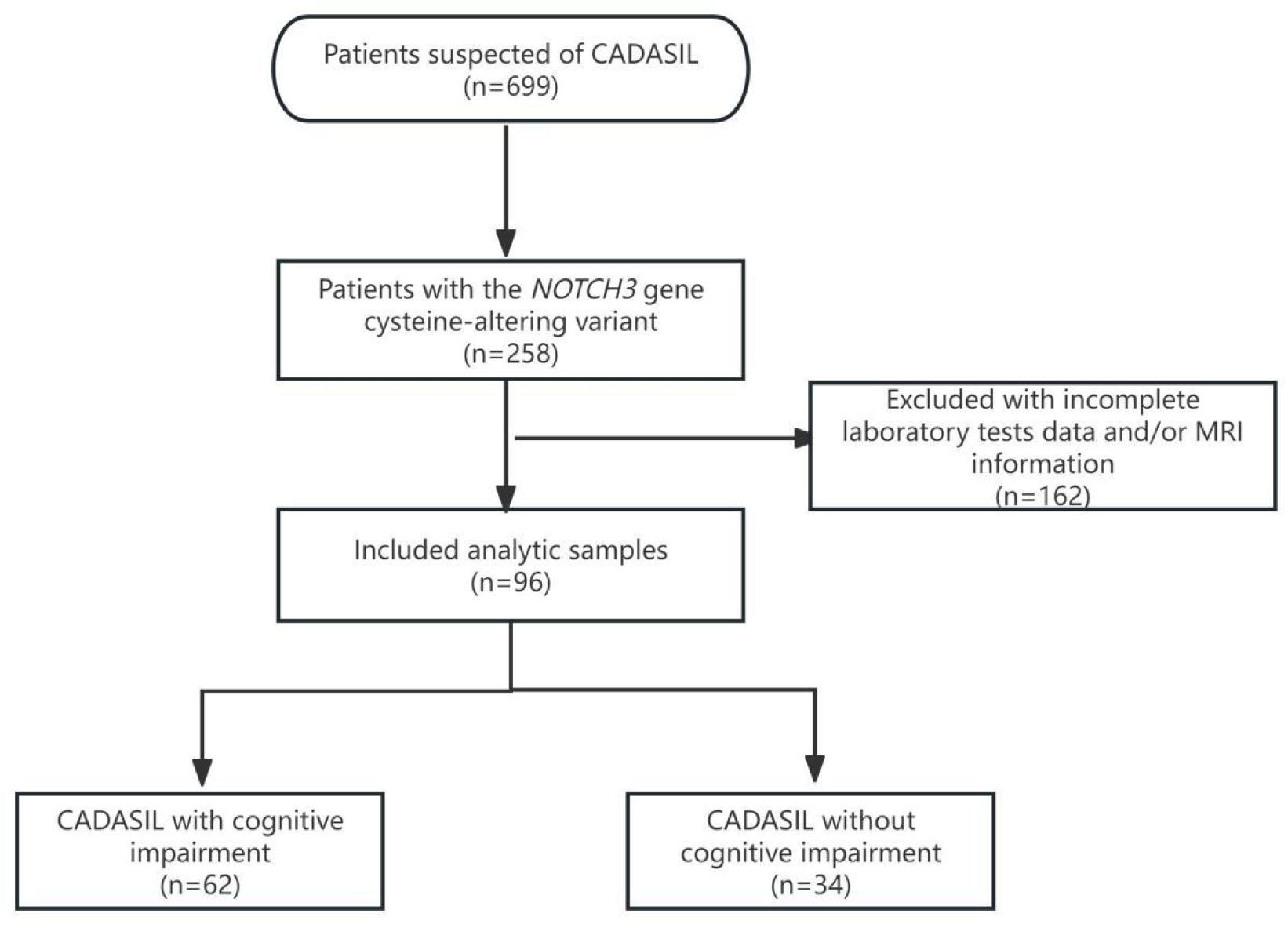
Flowchart of participant recruitment: CADASIL, Cerebral autosomal dominant arteriopathy with subcortical infarcts and leukoencephalopathy; MRI, magnetic resonance imaging

### 2.2 Clinical characteristics

#### Baseline clinical assessment and Laboratory measurement

Clinical characteristics included Baseline subject demographic characteristics, medical history (hypertension, hyperlipidemia, diabetes), risk factors (including gender, age, years of education, drinking history, smoking history) documented at enrollment.

All participants accepted a Mini-Mental State Examination (MMSE) assessment and venous blood routine laboratory tests. Cognitive impairment was defined as MMSE score <27 at baseline. Flow cytometry measurement of neutrophil, platelet, and lymphocyte counts (Machine model, XE-2100, SYSMES, Japan), SII = neutrophil count(×10^9^/L) × platelet count (×10^9^/L) / lymphocyte count(×10^9^/L).

### 2.3 MRI acquisition and evaluation

All patients underwent brain scanning with the same standardized 3.0T MRI protocol. Brain MRI scans was performed using 3D T1-weighted, T2-weighted, and FLAIR sequences, as well as either T2* or susceptibility-weighted imaging (SWI). Image data were analyzed by hospital trained personnel from the First Affiliated Hospital of Fujian Medical University. Radiographic assessment of each CSVD image marker was scored by two experienced doctors according to the standards for reporting vascular changes on neuroimaging (STRIVE), If the results was inconsistent, another senior doctor will conduct a re-evaluation. All physicians were unaware of the participants’ clinical data.

Lacune were delimited as subcortical oval or round lesions of CSF signal measuring 3mm to 20mm in diameter on T2WI and FLAIR images. The severity of lacunae is classified according to the total number of lacunae: mild (1-3), moderate (4-10), and severe (>10). Deep white matter hyperintensities (WMH) and periventricular appear as increased brightness on FLAIR and T2-weighted images. WMH were assessed using the Fazekas scale (range 0–3). Severe white matter hyperintensities: pWMH score =3 or dWMH score ≥2. White matter hyperintensities (WMH) are defined as increased brightness on fluid-attenuated inversion recovery (FLAIR) and T2-weighted imaging (T2WI) images. WMH are assessed using the Fazekas scoring scale (0 to 3 points). Severe WMH are defined as a deep white matter hyperintensities(dWMH) score of 2 or 3 and a periventricular white matter hyperintensities (pWMH) score of 3. Cerebral microbleeds (CMBs) appear as round, hypodense lesions with a diameter ranging from 2 to 10 mm on T2* or sensitivity weighted images (SWI). CMBs are classificated based on their total count: Grade 1 (1-8) and Grade 2 (≥9). On T2-weighted images, enlarged perivascular spaces (EPVS) appear as small punctate or linear hyperintensities measuring less than 3 mm in diameter in the basal ganglia and centrum semiovale. We used a validated 4-point visual rating scale to assess EPVS (0: none, 1: 1-10 EPVS, 2: 11-20 EPVS, 3: 21-40 EPVS, 4: >40 EPVS), with severe EPVS defined as a score ≥2.According to the Wardlaw group scoring system, The SVD scoring scale ranges from 0 to 4: (1)dWMH: Fazekas score ≥2 or pWMH: Fazekas score = 3, (2) lacuna: presence of one or more lacunae, (3) CMBs: presence of one or more CMBs, (4) EPVS: EPVS score ≥2. We considered this to be a severe cerebral small vessel disease burden when the score exceeded 2.

### 2.4 Statistical analysis

Continuous variables that are not normally distributed are presented as median with interquartile range; Continuous variables that follow a normal distribution are typically expressed as the mean with the standard deviation. Non-parametric test (Mann-Whitney U test) or two-tailed Student’s T test was employed for comparing differences. Categorical variables are expressed in terms of frequency. Chi-square test, Chi-square test with continuity correction, or Fisher’s exact test was used for comparisons of categorical variables between the two groups. The relationship of SII and cognitive impairment was evaluated. Develop a logistic regression model for cognitive impairment in patients with CADASIL, and calculate the odds ratios (OR) with their 95% confidence intervals (CI). Each outcome were performed as two models. In Model 1: unadjusted; In Model 2: gender, age, and years of education were adjusted. The receiver operating characteristic (ROC) curve was constructed, the area under the curve (AUC) and the optimal cut-off point for SII levels by the Joden index were calculated to evaluated the model’s ability to predict cognitive impairment in patients with CADASIL. Spearman correlation analysis and ordered logistic regression analysis were performed on SII levels and summary SVD score or imaging marker scores. In all results, a p-value of less than 0.05 was considered statistically significant. Statistical analyses were performed using SPSS software (version 27.0; IBM, USA).

## 3 Results

### 3.1 Baseline characteristics of the Participants

At the baseline, the median age of 96 patients with CADASIL was 59.00 (52.25-66.75) years, and 56 (58.3%) individuals were male. Based on MMSE score, 62 participants with CADASIL presented with cognitive impairment, while 34 participants with CADASIL did not present. Participants with cognitive impairment were female, older, less educated, higher Nc, NLR, PLR, SII than those without cognitive impairment. Patients with cognitive impairment had higher proportions of severe lacunes, severe ventricular white matter hyperintensities, severe microbleeds, and higher SVD scores compared to those without cognitive impairment (*P*<0.05)(Figure 2). 96 patients and details of the two groups were detailed (Table 1).

**Figure 2.**
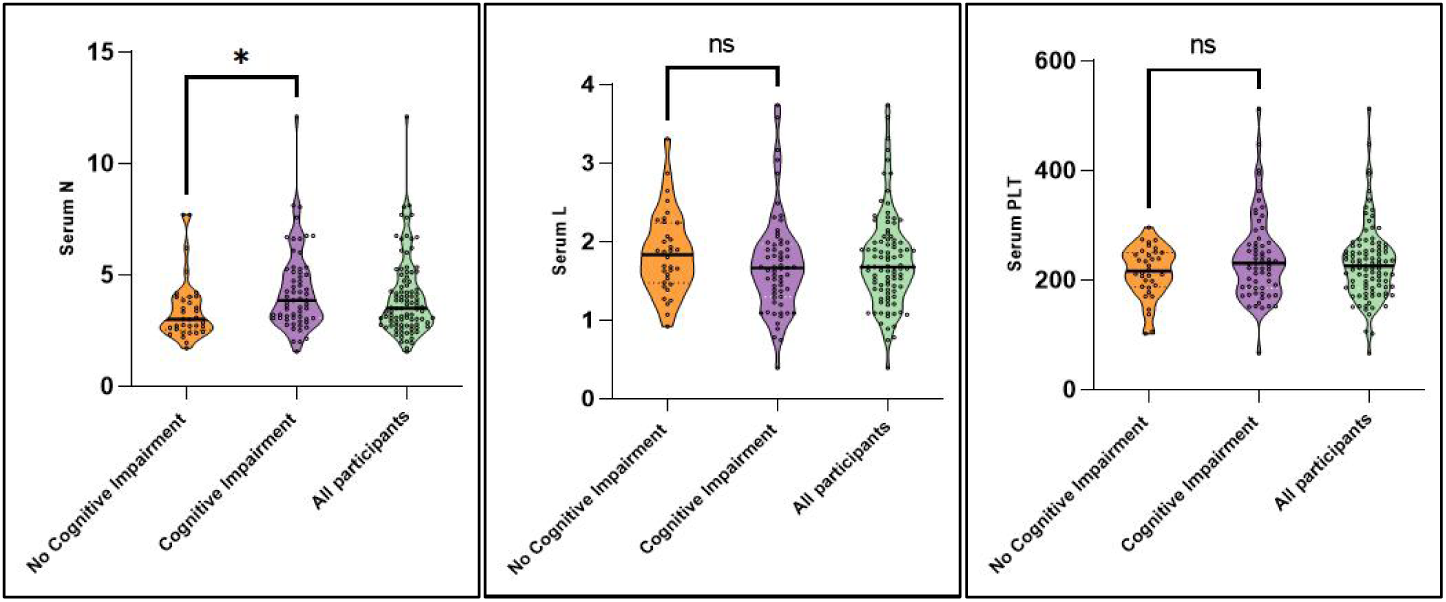
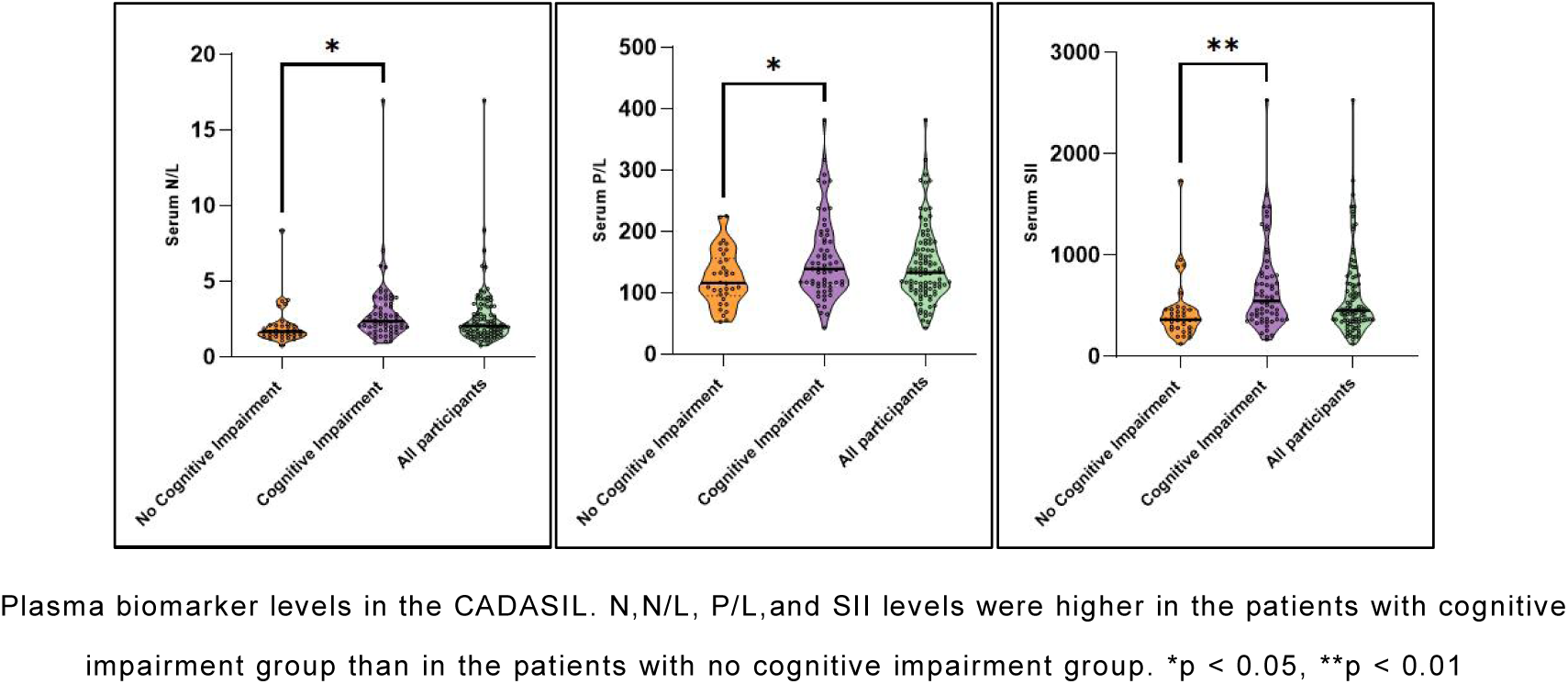
Plasma biomarker levels in the CADASIL Between Cognitive Impairment Group and No Cognitive Impairment Group

**Table 1.**
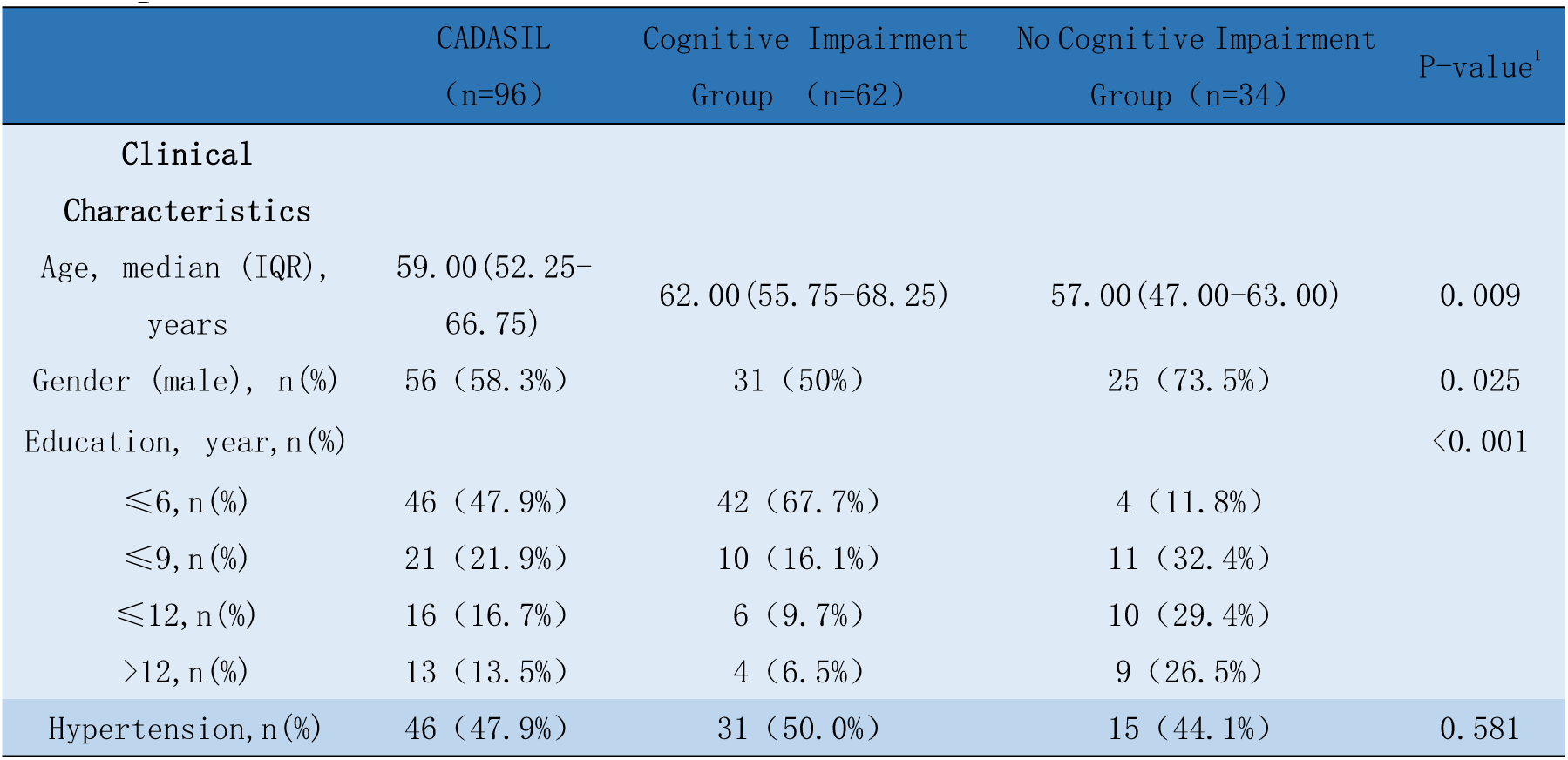
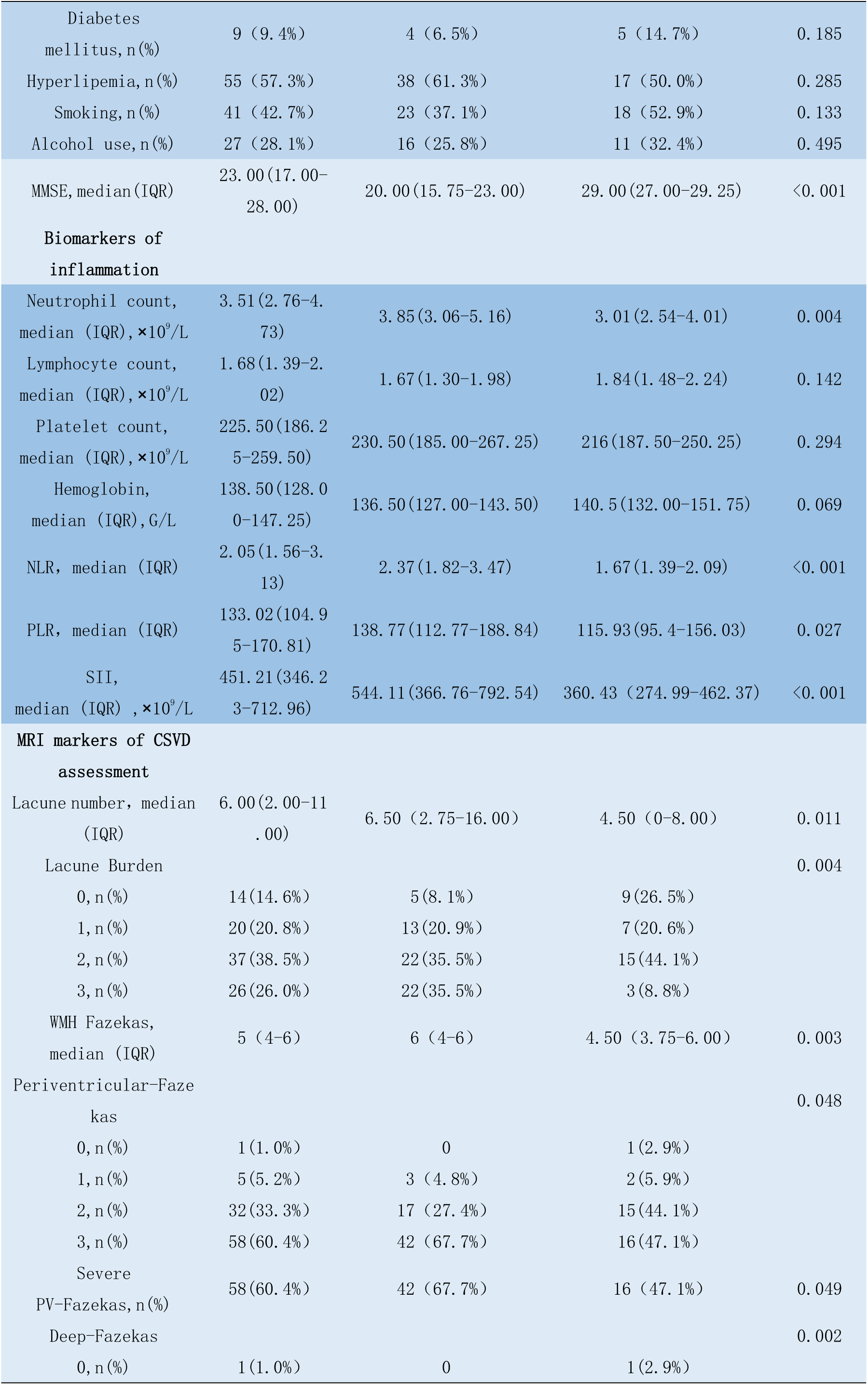
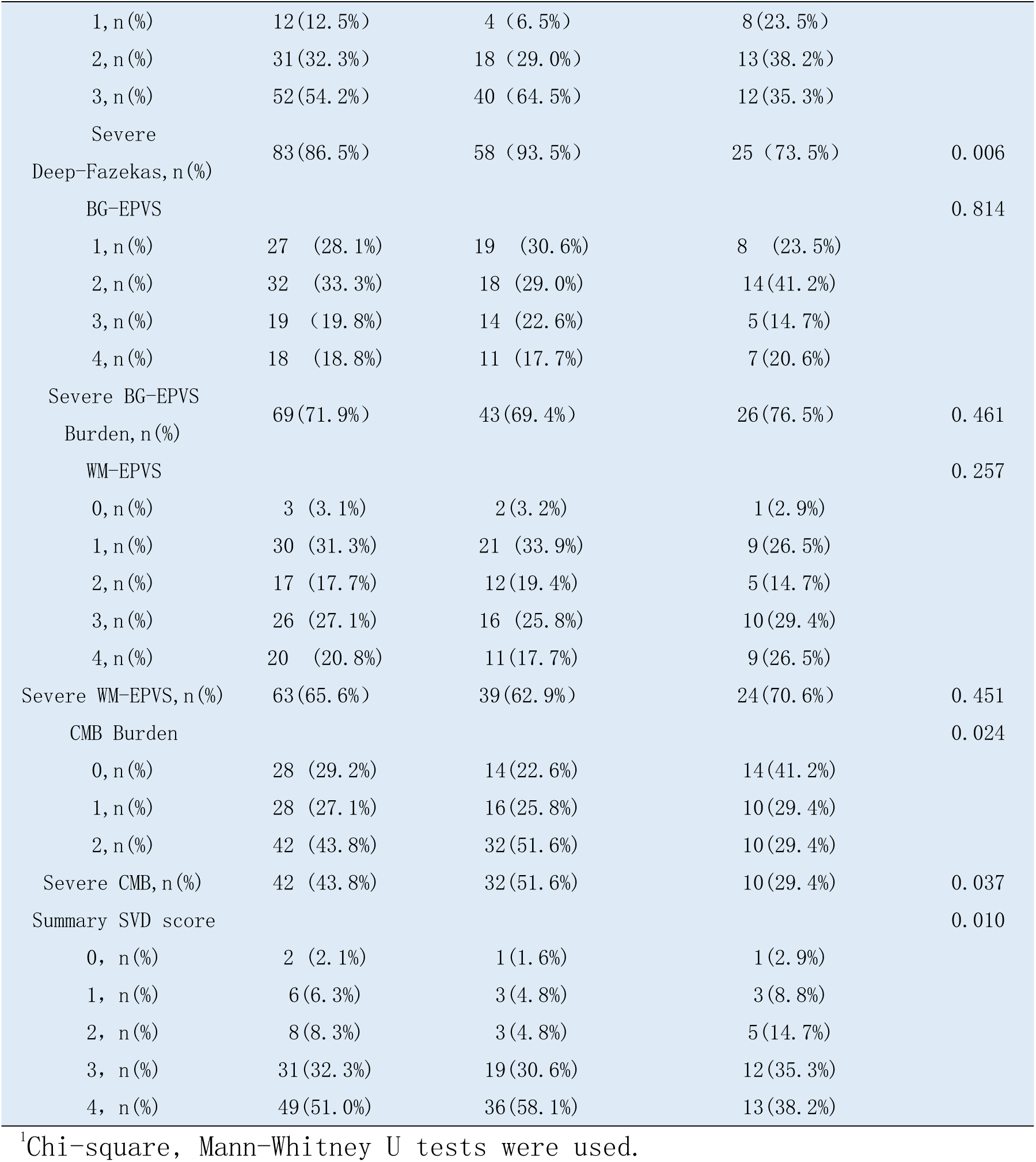
Baseline Clinical Characteristics and MRI markers of CADASIL Between Cognitive Impairment Group and No Cognitive Impairment Group.

### 3.2 Association between Cognitive Impairment and SII

The correlation between SII level and MMSE score in CADASIL patients was negatively correlated by Spearman correlation analysis (r=-0.336, *P*<0.001)(Figure 3). ROC curve for SII to identify cognitive impairment in CADASIL patients: AUC area under curve 0.713 (*P*<0.001), SII optimal cut-off value 495.854, sensitivity 0.548, specificity 0.853 (Figure 4). Unadjusted binary logistic regression showed that patients in the last quartile (Q4) of SII were linked with a higher risk of developing cognitive impairment than those in the first quartile (Q1)(Q4 vs. Q1: OR 5.909, 95% CI 1.546 -22.580, *P*=0.009); After resetting for gender, age and education, the last quartile (Q4) of SII was also suggestively associated with developing cognitive impairment.(Q4 vs. Q1: OR 5.230, 95% CI 1.040 -26.297, *P*=0.045)(Table 2).

**Figure 3.**
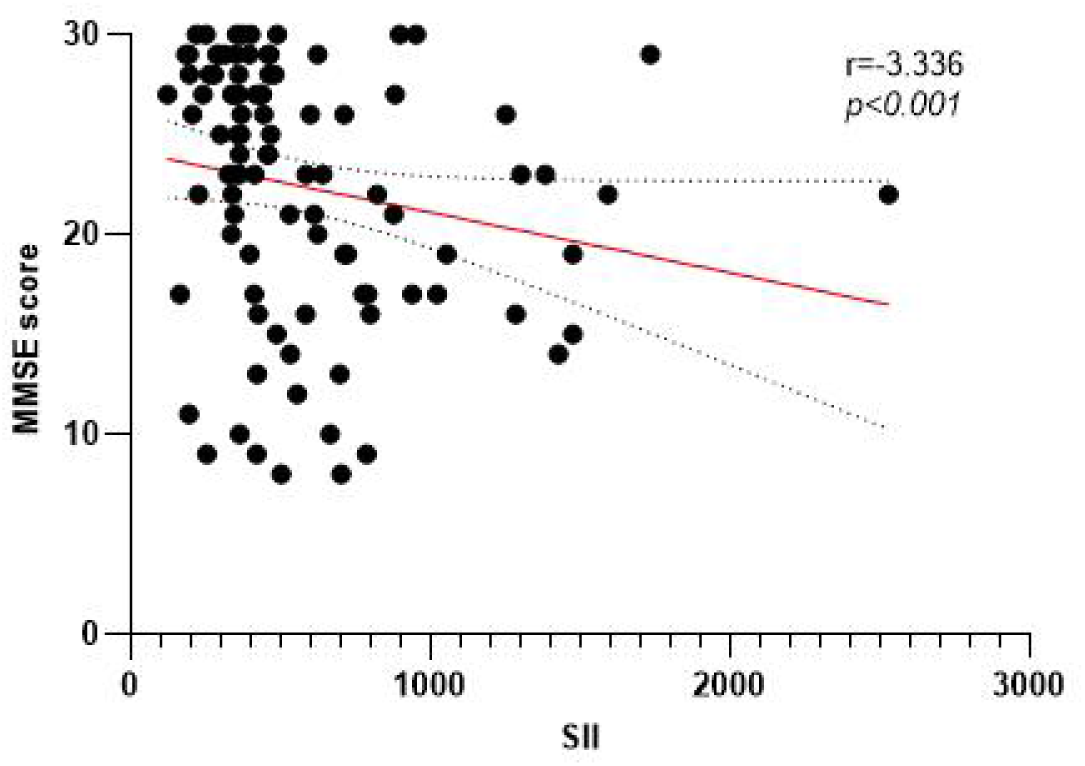
Scatterplot of SII and MMSE score.

**Figure 4.**
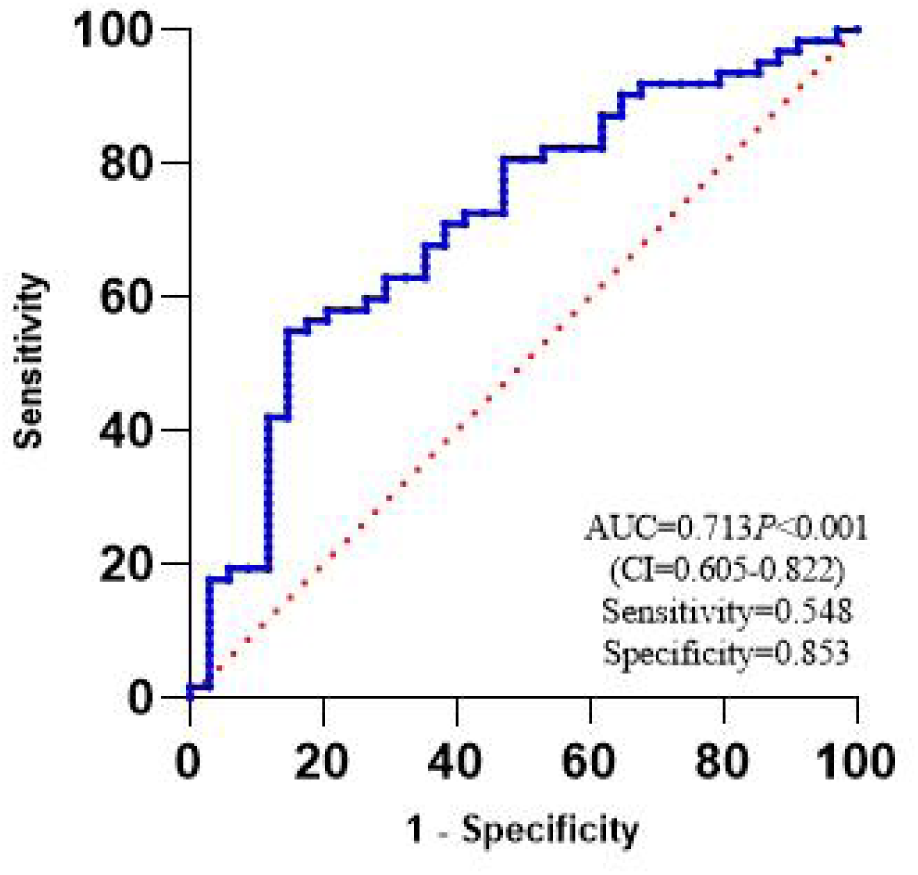
Receiver operating characteristic (ROC) curve of SII levels for cognitive impairment.

**Table 2.**
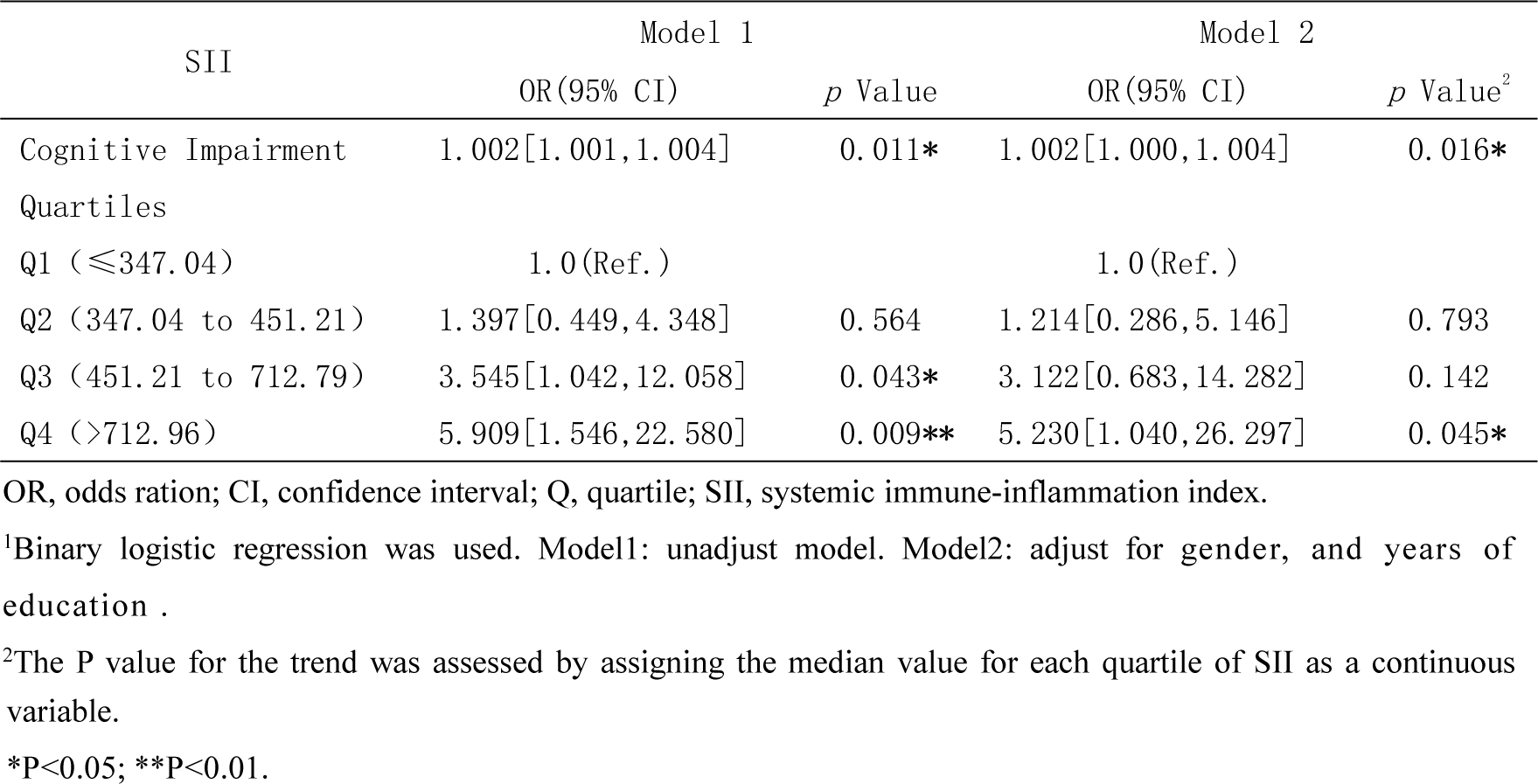
Association between SII and Cognitive Impairment^1^.

### 3.3 Correlation between SII and MRI CSVD imaging markers and summary SVD score

In this study, we performed Spearman correlation analysis between SII levels and summary SVD score and MRI imaging marker scores in CADASIL patients. Unexpectedly, There was no noticeable difference between SII and MRI imaging marker scores, nor with SVD scores (Figure 5). A multivariate logistic regression model was used to investigate the associations between SII, CSVD, neuroimaging markers, and the total SVD score. There was no significant correlation between SII levels and MRI imaging markers, including white matter hyperintensity score, paraventricular white matter hyperintensity score, semi-oval center white matter hyperintensity score, lacunar lesion score, semi-oval center EPVS score, basal ganglia EPVS score, and microhemorrhage score (Figure 6).

**Figure 5.**
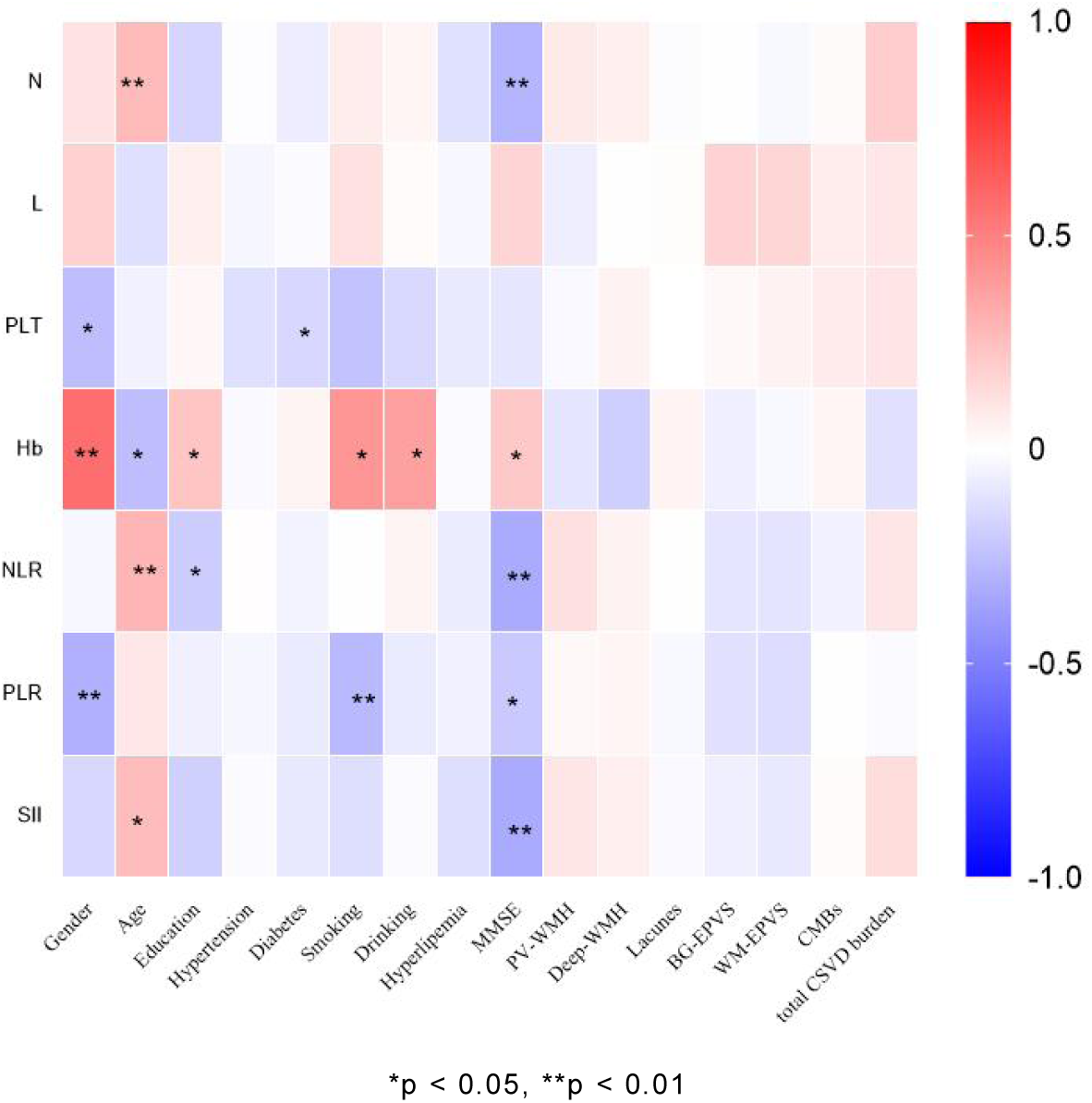
Spearman Correlation Analysis of SII and MRI imaging markers of CSVD

**Figure 6.**
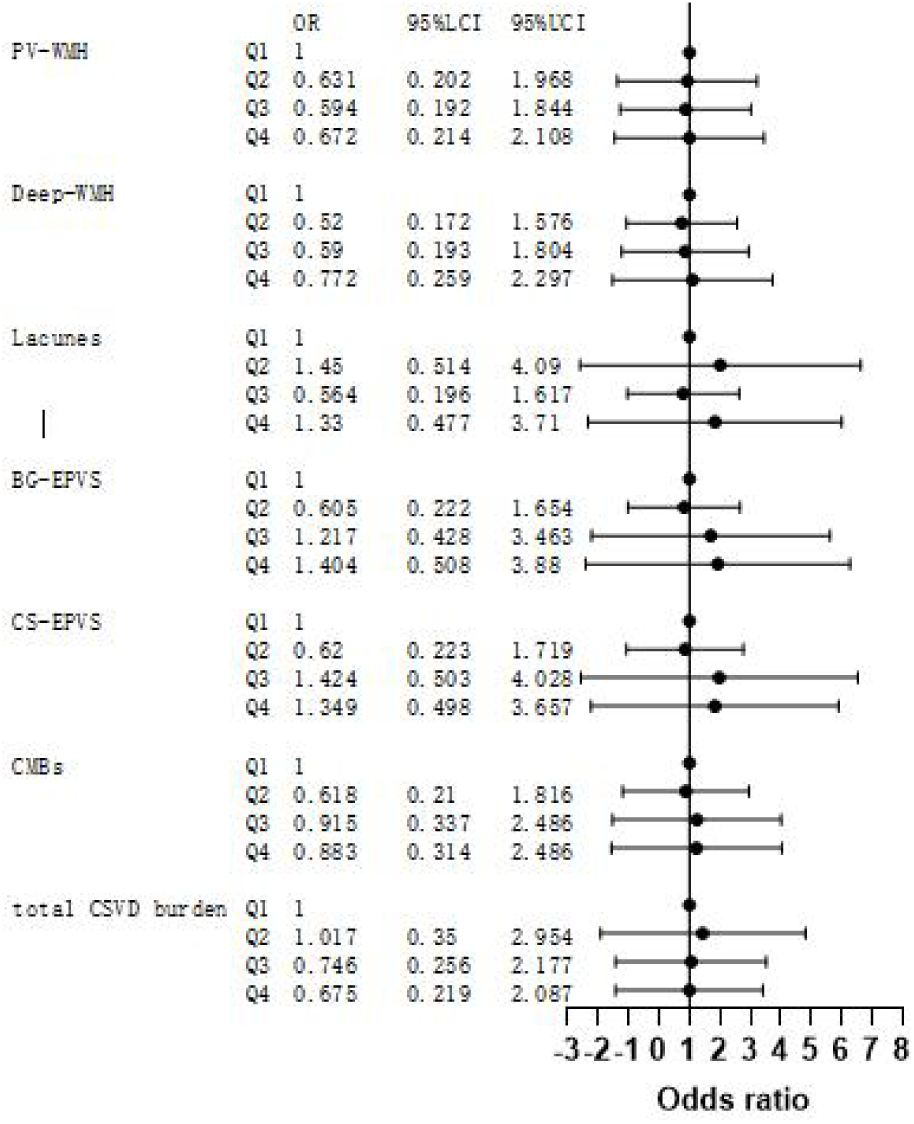
Forest plots for the association of SII and MRI imaging markers or total CSVD burden.

## 4 Discussion

The SII is a novel inflammatory measure that has emerged in recent years, defined as the platelet count × neutrophil count / lymphocyte count in a peripheral venous blood sample^(17)^. SII was originally identified as a cancer patient with a high risk of mortality or recurrence, who could benefit from early intervention^(13)^. Recently, the SII index has been shown to predict functional outcomes in acute ischemic stroke, cerebral hemorrhage, and subarachnoid hemorrhage^(14–21)^, and in patients with CADASIL, SII continues to be a novel systemic inflammatory biomarker that has not been fully explored. In this study, we found that patients without cognitive impairment had lower SII levels than CADASIL patients with cognitive impairment, and an increase in SII was positively linked with the risk of developing cognitive impairment. ROC curve analysis showed that SII could predict cognitive impairment in patients with CADASIL (AUC=0.713 *P*=0.001), SII optimal cut-off value 495.854, sensitivity 0.548, specificity 0.853, suggesting that SII value may be a promising predictive biomarker for cognitive impairment in CADASIL. SII value ≥495.854 at baseline may be a marker to guide clinicians to screen high risk patients with cognitive impairment in CADASIL. The logistic regression results suggested that SII could be used to predict cognitive impairment in CADASIL patients, and elevated SII may be an independent risk factor for cognitive impairment in patients with CADASIL. Concerning clinical management of cognitive impairment in CADASIL patients, we can early detect and stratify the risk in patients with CADASIL according to SII level, which may be helpful for early detection of potential cognitive impairment symptoms, so as to adjust treatment regimens in time, so as to better individualize management and treatment. In recent years, multiple studies have suggested a potential correlation between SII and cognitive impairment in different diseases, this aligns with our observations. A retrospective research found that patients with elevated SII are more prone to postoperative cognitive impairment among elderly patients^(22)^. A studies have found that there is an independent correlation between SII and cognitive impairment after stroke in patients at admission. Higher SII level may be an effective predictor of cognitive impairment after stroke^(23)^. A cross-sectional analysis included 2,194 older adults participants suggested a positive association between cognitive function and SII^(24)^. Evidence from other studies also supports a strong connection between SII and cognitive impairment^(25–27)^. However, the underlying mechanism of this association remains unclear. Current research has demonstrated that inflammation plays a critical role in the onset and progression of CSVD. Systemic inflammatory processes were believed to be linked with endothelial dysfunction, cerebral blood flow autoregulation and blood-brain barrier permeability, which might impact on the occurrence and progression of cerebral small vessel disease^(12)^.

Our study also revealed there was no significant connection between SII and CSVD imaging markers or summary SVD score in CADASIL. Previous correlations between SII and CSVD imaging markers and summary SVD score were mixed. summary SVD scores were previously reported to be significantly negatively correlated with SII levels in CSVD^(28)^. In healthy individuals, higher SII levels are linked with a greater volume of WMH^(29)^. Studies based on cross-sectional and longitudinal data including from the UK Biobank cohort suggest that SII and neutrophil/lymphocyte ratio may be early indicators of white lesions in CSVD patients, suggesting that systemic inflammatory response may be involved in the mechanism of early white lesions in CSVD patients^(30)^. A community-based prospective cohort study shows that higher SII levels were positively correlated with WMH scores and moderate to severe BG-EPVS, however no significant correlation was observed between SII and summary SVD score^(12)^. Our study did not suggest that there was a strong correlation between SII and CSVD imaging markers or summary SVD score in CADASIL patients. The conclusions of our study were not completely consistent with previous related studies. It might be related to the higher proportion of imaging markers and severe summary SVD score in CADASIL patients compared with sporadic small vessel disease patients at diagnosis in previous related studies.

This CADASIL study reports novel links between SII, cognitive impairment, and CSVD imaging; however, our study is not without its limitations. First, due to the cross-sectional study design, it is difficult to obtain a clear causal relationship between SII and cognitive impairment or CSVD imaging, and further prospective studies are needed. Secondly, this study excluded patients with missing clinical data, blood routine or magnetic resonance imaging values, which may cause selection bias to a certain extent. The participants in this study were from southern China, which may cause a certain degree of selection bias. The relatively small sample size of our study necessitates future validation in larger, multicenter cohorts. Third, our analysis adjusted for several potential confounders, however we could not completely exclude unmeasured covariates. Fourth, MMSE did not adequately represent the complexity of certain cognitive domains and required the use of additional cognitive screening tools. These issues need to be addressed in further follow-up studies.

Overall, this study found that elevated SII levels in CADASIL patients are associated with cognitive impairment. Nevertheless, there were no statistically noteworthy differences between SII and summary SVD score or imaging markers. SII provides a reliable, objective, specific, and noninvasive method for potentially predicting the onset of cognitive impairment, and provides clinical guidance for screening, treatment, and prognostic assessment of cognitive impairment, which is an area worthy of further investigation. However, the causal relationship of this association needs to be further confirmed through multi-center, prospective, large-sample studies.

## Abbreviations

SII: immune-inflammation index
NLR: neutrophil-to-lymphocyte ratio
PLR: platelet-to-lymphocyte ratio
OR: odd ratio
CSVD: Cerebral small vessel disease
CADASIL: Cerebral autosomal dominant arteriopathy with subcortical infarcts and leukoencephalopathy
MMSE: Mini-Mental State Examination

## Acknowledgments

We would like to thank all participants and their families for their Cooperation. We are also grateful to the staff.

## Author Contributions

JHP: writing – Original draft, Data curation, Visualization, Methodology, Investigation. HRZ: Conceptualization, Data curation, Writing – review and editing. FWH: Data curation, Investigation. GPL: Data curation, Investigation. ZP: Investigation, Data curation. SYX: Investigation, Data curation. TY: Investigation, Data curation. WXQ: Investigation, Data curation. CHC: Investigation, Data curation. BC: Conceptualization, Methodology, Supervision, Resources, Software, Writing – review and editing.

## Funding

The author(s) declare that financial support was received for the research, authorship, and/or publication of this article. This study was supported by grant No. 82471299 (B.C.) and 82071277 (B.C.) from the Natural Science Foundation of China, grant No. 2023Y9039(B.C.) and 2018Y9084 (B.C.) Joint Funds for the Innovation of Science and Technology, Fujian Province, grant No. LHGJ20240434(J.P.) Joint Construction Project of Henan Medical Science and Technology Research and Development Program, Henan Province.

## Data Availability Statement

The data that support the findings of this study are available from the corresponding author upon reasonable request. The data are not publicly available due to privacy or ethical restrictions.

## Declarations

### Ethical approval and consent to participate

The study was approved by the Ethics Committee of the First Affiliated Hospital of Fujian Medical University, China (MRCTA, ECFAH of FMU(2019)245). The study was conducted in accordance with the principles of the Declaration of Helsinki. Written informed consent was obtained from the individual(s), and legal guardian/next of kin.

### Consent for publication

Not applicable.

### Competing interests

The authors declare there are no conflicts of interest – financial or otherwise – related to the material presented herein.

## Notes

### Competing Interest Statement

The authors have declared no competing interest.

### Clinical Trial

ClinicalTrials.gov identifier: (NCT04318119)

### Funding Statement

Yes

### Author Declarations

The study was approved by the Ethics Committee of the First Affiliated Hospital of Fujian Medical University,China(MRCTA,ECFAH of FMU(2019)245). The study was conducted in accordance with the principles of the Declaration of Helsinki.The studies were conducted in accordance with the local legislation and institutional requirements. Written informed consent was obtained from the individual(s), and legal guardian/next of kin, for the publication of any potentially identifiable images or data included in this article.

